# Maximising information on smell, quantitative motor impairment and probable REM-sleep behaviour disorder in the prediction of Parkinson’s disease

**DOI:** 10.1101/2020.03.03.20023994

**Authors:** Jonathan P. Bestwick, Stephen D. Auger, Anette E. Schrag, Donald G Grosset, Sofia Kanavou, Gavin Giovannoni, Andrew J. Lees, Jack Cuzick, Alastair J. Noyce

**Author notes:** **Correspondence to:** Jonathan Bestwick, Preventive Neurology Unit, Wolfson Institute of Preventive Medicine, Queen Mary University of London, Charterhouse Square, London, EC1M 6BQ, UK. **Funding agencies:** Both the PREDICT-PD and Tracking Parkinson’s studies are funded by Parkinson’s UK. **Relevant conflicts of interests/financial disclosures:** Nothing to report.

## Abstract

**Background:** Hyposmia, motor impairment and probable REM-sleep behaviour disorder (RBD) are markers for Parkinson’s disease (PD). Proposed PD risk prediction models have dichotomised test results and applied likelihood ratios (LRs) to scores above and below cut-offs. We investigate whether LRs for specific test values could enhance prediction models.

**Methods:** Smell and probable RBD data for PD patients were taken from the *Tracking Parkinson’s* study (n=1046). For motor impairment previously published data were supplemented (n=87). PREDICT-PD pilot study participants were the controls. Smell, motor impairment and RBD were assessed using the University of Pennsylvania Smell Identification Test (UPSIT), the Bradykinesia-Akinesia Incoordination (BRAIN) test and the REM sleep behaviour disorder Screening Questionnaire (RBDSQ). UPSIT and RBDSQ data were analysed using logistic regression to determine which items were predictive of PD, or using total scores. Gaussian distributions were fitted to BRAIN test scores. LRs were calculated from logistic regression models or from score distributions. False-positive rates (FPRs) for specified detection rates (DRs) were calculated.

**Results:** Logistic regression modelling yielded a greater range of LRs. 16 odours were associated with PD; LRs ranged from 0.005-5511. 6 RBDSQ questions were associated with PD; LRs ranged from from 0.34-69. BRAIN test LRs ranged from 0.16-1311. For a 70% DR the FPR for the 16 odours was 2.4%, for 50% DRs, the BRAIN test FPR was 6.6% and 12.2% for the RBDSQ.

**Conclusions:** Maximising information on PD markers can potentially improve the ability of algorithms to detect PD by generating LRs with a larger range of values than using dichotomised results.

## INTRODUCTION

Parkinson’s disease (PD) affects about 1% of individuals over the age of 60.^1^ Clinical PD diagnosis is usually made late in the disease process and current treatments only relieve symptoms. Identifying earlier stages of PD may increase our chances of slowing disease progression.^2,3^ Accordingly, risk prediction models have been developed. In PREDICT-PD, a pilot study of 1323 individuals aged 60-80 recruited from the general UK population^4^, the risk of PD was estimated based on a systematic review and meta-analysis of risk factors and early features.^5^ Separately, the Movement Disorders Society (MDS) produced criteria for the diagnosis of prodromal PD^6,7^, a risk algorithm based on primary care presentations has been described^8^ and a risk algorithm based on clinical and genetic classification.^9^

In reporting the baseline and year 3 follow-up data from the PREDICT-PD pilot study, preliminary support for the validity and the value of the risk algorithm was assessed by comparing ‘intermediate markers’ for PD between those at estimated higher and lower risk. These intermediate markers included three of the strongest indicators of increased PD risk: hyposmia, reduced finger tapping speed, and probable REM-sleep behaviour disorder (RBD).^4,10^ REM sleep behaviour disorder was assessed subjectively using the REM sleep behaviour disorder Screening Questionnaire (RBDSQ), with a score of ≥5 indicating probable RBD.^11^

Hyposmia was assessed objectively using the University of Pennsylvania Smell Identification Test (UPSIT), a 40-item “scratch-and-sniff” smell test.^12^ A score ≤15^th^ centile was used to indicate hyposmia, which equated to an UPSIT score of ≤27.^4,10^ Recently, members of our group used a data-driven approach to propose smaller subsets of the 40 items in the UPSIT which could be used on a wider scale to predict hyposmia.^13^ *Finger tapping speed* was used as a quantitative motor marker with the Bradykinesia Akinesia Incoordination (BRAIN) test.^14^ Users completed the BRAIN test online by alternately tapping the ‘S’ and ‘:’ keys on a keyboard as rapidly and accurately as possible in 30 seconds. The two most useful parameters generated were the kinesia score (KS, the total number of key taps) and the akinesia time (AT, the mean dwell time of keys in milliseconds). In the three-year follow-up of PREDICT-PD, a KS score ≤44 (≤15^th^ centile) was used to indicate reduced tapping speed.^10^

Given that smell loss, reduced tapping speed and RBD are all recognised markers for PD, combining them in a risk prediction algorithm would be expected to improve risk estimation, improving discrimination between those that develop the disease and those who do not.

Common to all PD-prediction models is the modification of the age-related risk of PD using the presence or absence of risk factors or protective factors. The MDS criteria does this using likelihood ratios: e.g. the likelihood ratios for those with hyposmia is 6.4 and 0.40 for those without; for probable RBD (based on the RBDSQ) the likelihood ratios (with and without) are 2.8 and 0.89, and for abnormal quantitative motor testing the likelihood ratios (with and without) are 3.5 and 0.60.^7^ The approach of dichotomising tests results loses information for these continuous or discrete markers.^15^ Here, we investigated whether the use of likelihood ratios generated from the full range of values rather than dichotomised values yields a greater spread of risk, and hence an expected better risk prediction model.

## METHODS

### Data sources

We used data from individuals in the PREDICT-PD pilot study who had not had a diagnosis of PD during follow-up (mean age 67 years, 62% female) who had either completed the UPSIT and/or completed the BRAIN test and/or completed the RBDSQ. Full details of the study were previously published.^4^ Data from cases with PD were derived from several sources: the assessment of UPSIT and RBDSQ scores were taken from baseline data of those aged between 60 to 80 years in the *Tracking Parkinson’*s study, a multicentre prospective longitudinal epidemiological and biomarker study of PD (n=1046, mean age 69, 53% male; RBDSQ scores available on n=983).^16^ For the BRAIN test, 59 PD cases came from published data^17^, supplemented with unpublished data for a total of n=87 patients with PD. BRAIN tests were performed ‘off’ treatment in PD patients.

### Calculation of continuous scores

For the full 40-item UPSIT, scores were adjusted by performing a median regression of UPSIT scores against age and gender among controls, then subtracting all participants’ UPSIT scores from their expected score from the regression equations (UPSIT delta values). To investigate whether there was a mixture of Gaussian distributions among PD patients and controls, finite mixture models with 2 Gaussian components were fitted. From the final distributions, likelihood ratios according to UPSIT delta values were calculated as the height of the modelled distribution (density) in PD patients divided by the height of the modelled distribution in controls. To avoid the phenomena of risk reversal, which occurs when the standard deviation of a screening marker in an affected population is greater than that in an unaffected population, truncation limits were applied so that the likelihood ratio is a monotonically increasing function of UPSIT delta values.^18^ In addition to using the full 40-item UPSIT, we also performed logistic regression to determine which odours were predictive of PD. A forward stepwise procedure was used for this analysis (with a 0.05 significance level for entry into the model), using the 32 odours that were common to both the UK and US version of the UPSIT (The PREDICT-PD pilot used the US version while the Tracking Parkinson’s study used the UK version). We also explored the performance of a subset of the 6 odours most strongly associated with PD identified from the forward stepwise procedure. We further examined the results of a 6-item smell test which was based on the four odours that discriminated most between those with and without hyposmia from previous work, plus a further two odours that most discriminated between PD and controls (menthol, clove, orange, onion, coconut and cherry).^13,19^

For the BRAIN test, results for the worst of KS and AT scores from each hand were adjusted by performing a median regression of scores against age and gender among controls, then either subtracting (for KS) or dividing (for AT) all participants’ scores by the expected score from the regression equations. Fits of adjusted BRAIN test scores to Gaussian distributions were assessed by inspection of probability plots. The points at which data started to deviate from a Gaussian distribution were used as truncation limits unless there were any point of risk reversal within the limit, in which case the point of risk reversal was used as a truncation limit. The means, standard deviations and correlation coefficients between the two parameters were calculated to define a bivariate Gaussian distribution in PD patients and controls. To avoid the influence of outliers, the median was used as the mean and robust standard deviations were calculated as the 90^th^ centile minus the 10^th^ centile divided by 2.563 (i.e. number of standard deviations between the 10^th^ and 90^th^ centiles of a Gaussian distribution). Likelihood ratios according to adjusted KS and AT values were calculated as the height of the bivariate Gaussian distribution in PD patients, divided by the height of the bivariate Gaussian distribution in controls.

For total RBDSQ scores, the final question which scores 1 if a person was diagnosed with a disease of the nervous system was not included, given that all PD patients would score 1. The likelihood ratio for each score (from 0 to 12) was calculated as the percentage of PD patients with that score, divided by the percentage of controls who scored the same. A log-quadratic regression (weighted by the total number of PD patients and controls with each score) was performed to obtain a ‘smooth’ function for the likelihood ratio according to score. As with analyses on UPSIT scores, logistic regression was also performed to determine which questions in the RBDSQ were predictive of PD and the two approaches were compared.

For each intermediate marker and approach, the performance in predicting PD was estimated as false-positive rates for specified detection rates. The area under the receiver operating characteristic (ROC) curve was also calculated (AUC). Internal validation of the specified models was performed using the bootstrapping method.^20^ Briefly, for the fitted logistic regression models using all data, a model using the same variables (odours, RBDSQ questions) was fitted on a bootstrap sample, and that model tested on the bootstrap sample and on the original data, with the AUC and false-positive rates for specified detection rates calculated. This process was repeated 1000 times. The average difference in the AUC, and false-positive rates for specified detection rates provided estimates of the optimism of the performance of the models fitted on all data. The estimates of optimism were then subtracted from the performance measures to estimate the internally validated performance. For the BRAIN test a similar method was used, but based on fitting bivariate Gaussian distributions to bootstrap samples. Statistical significance was set at 5% and all analyses were performed using Stata version 15 (StataCorp, College Station, Texas).

### Ethical approval

The PREDICT-PD study was approved by Central London Research Committee 3 (reference number 10/H0716/85). 72 sites in the UK providing secondary care treatment for PD patients as part of the UK National Health Service (NHS) (and in selected sites, their linked academic institutions) are participating in the Tracking Parkinson’s study, with multicentre ethics committee and local research and development department approvals.

## RESULTS

### Smell

The median number of correctly identified odours (out of 40) was 18 among PD patients and 32 among controls (p<0.001). Among the 32 odours common to both the UK and US versions of the UPSIT, correlation coefficients between pairs of odours were low; all less than 0.3, with 95% of correlations less than 0.2. Table 1 shows the results of the multivariate logistic regression analyses on items from the UPSIT. From the 32 odours, 16 were found to be significantly associated with PD in a multivariate model (Supplementary Table 1 shows univariate odds ratios for each of the 32 odours). The table also shows the results for the 6 odours that were most strongly associated with PD. Likelihood ratios are calculated by using the coefficients in Table 1 to generate the log odds, exponentiating and then dividing by 932/887; the number of PD cases divided by the number of controls in the analysis). Figure 1 shows the distribution of likelihood ratios and ROC curves for each set of odours. Based on the 16 odours the median likelihood ratios for PD were 36 and 0.05 in PD patients and controls respectively (p<0.001) and ranged from 0.009 to 5511 in PD patients and from 0.005 to 515 in controls. False-positive rates for 50, 60, 70 and 80% detection rates were 1.1%, 1.9%, 2.4% and 3.7% respectively. Corresponding internally validated estimates were 1.2%, 2.1%, 2.5% and 4.0%. The AUC was 0.97 and the internally validated AUC was 0.96. Based on the 6 odours that were most strongly associated with PD in this analysis, the median likelihood ratios were 32 in PD patients and 0.03 in controls (p<0.001) and ranged from 0.03 to 1429 in PD patients and from 0.03 to 315 in controls, and corresponding false-positive rates were 0.9%, 1.6%, 2.8% and 4.4% respectively. Corresponding internally validated estimates were 0.9%, 1.7%, 2.9% and 4.4%. The AUC was 0.95 and the internally validated AUC was also 0.95.

**Table 1:** Results of multivariate logistic regression analyses of the odours common to the UK and US versions of the UPSIT, and the 6 odours most strongly associated with Parkinson’s disease

| Odour | Model including all significant odours |  |  | Model including 6 odours most strongly associated with PD |  |  | Likelihood ratio test statistics between nested models |  |
| --- | --- | --- | --- | --- | --- | --- | --- | --- |
| | Coefficient | OR (95% CI) | p-value | Coefficient | OR (95% CI) | p-value | $\chi^2$ | p-value |
| Gasoline | -1.492617 | 0.22 (0.15 to 0.34) | $5.66 \times 10^{-12}$ | -1.952729 | 0.14 (0.1 to 0.21) | $2.17 \times 10^{-24}$ | 663.52 | $2.57 \times 10^{-146}$ |
| Soap | -1.956232 | 0.14 (0.10 to 0.21) | $4.43 \times 10^{-23}$ | -2.232339 | 0.11 (0.08 to 0.15) | $1.36 \times 10^{-34}$ | 354.48 | $4.486 \times 10^{-79}$ |
| Watermelon | -1.436059 | 0.24 (0.15 to 0.37) | $6.69 \times 10^{-11}$ | -1.859984 | 0.16 (0.11 to 0.23) | $5.82 \times 10^{-21}$ | 230.33 | $5.042 \times 10^{-52}$ |
| Lemon | -1.402295 | 0.25 (0.17 to 0.35) | $1.40 \times 10^{-14}$ | -1.511823 | 0.22 (0.16 to 0.3) | $6.21 \times 10^{-20}$ | 131.11 | $2.337 \times 10^{-30}$ |
| Cinnamon | -1.174883 | 0.31 (0.21 to 0.45) | $8.66 \times 10^{-10}$ | -1.544792 | 0.21 (0.15 to 0.3) | $2.35 \times 10^{-18}$ | 96.36 | $9.601 \times 10^{-23}$ |
| Natural gas | -1.084457 | 0.34 (0.21 to 0.54) | $6.15 \times 10^{-6}$ | -1.561454 | 0.21 (0.14 to 0.32) | $7.06 \times 10^{-13}$ | 55.57 | $9.03 \times 10^{-14}$ |
| Rose | -0.8652221 | 0.42 (0.28 to 0.63) | $2.71 \times 10^{-5}$ | | | | 38.76 | $4.79 \times 10^{-10}$ |
| Paint thinner | -0.8597572 | 0.42 (0.29 to 0.61) | $6.03 \times 10^{-6}$ | | | | 28.29 | $1.05 \times 10^{-7}$ |
| Pineapple | -0.7759867 | 0.46 (0.30 to 0.71) | 0.0004 | | | | 20.42 | $6.21 \times 10^{-6}$ |
| Banana | -0.6495947 | 0.52 (0.35 to 0.77) | 0.0011 |  |  |  | 13.07 | 0.0003 |
| Cedar | -0.5584326 | 0.57 (0.39 to 0.84) | 0.0048 |  |  |  | 10.25 | 0.0014 |
| Cherry | -0.5074133 | 0.60 (0.41 to 0.88) | 0.0087 |  |  |  | 7.12 | 0.0076 |
| Strawberry | 0.6116164 | 1.84 (1.21 to 2.81) | 0.0045 |  |  |  | 7.00 | 0.0082 |
| Coconut | -0.4896007 | 0.61 (0.41 to 0.91) | 0.0143 |  |  |  | 5.87 | 0.0154 |
| Menthol | -0.6320504 | 0.53 (0.32 to 0.88) | 0.0148 |  |  |  | 4.22 | 0.0400 |
| Mint | 0.5117308 | 1.67 (1.08 to 2.57) | 0.0209 |  |  |  | 5.48 | 0.0192 |
| Constant | 8.66404 | 5790.88 (2480.46 to 13519.39) | $3.09 \times 10^{-89}$ | 7.313897 | 1501.02 (754.9 to 2984.58) | $1.33 \times 10^{-96}$ | | |

**Figure 1:**
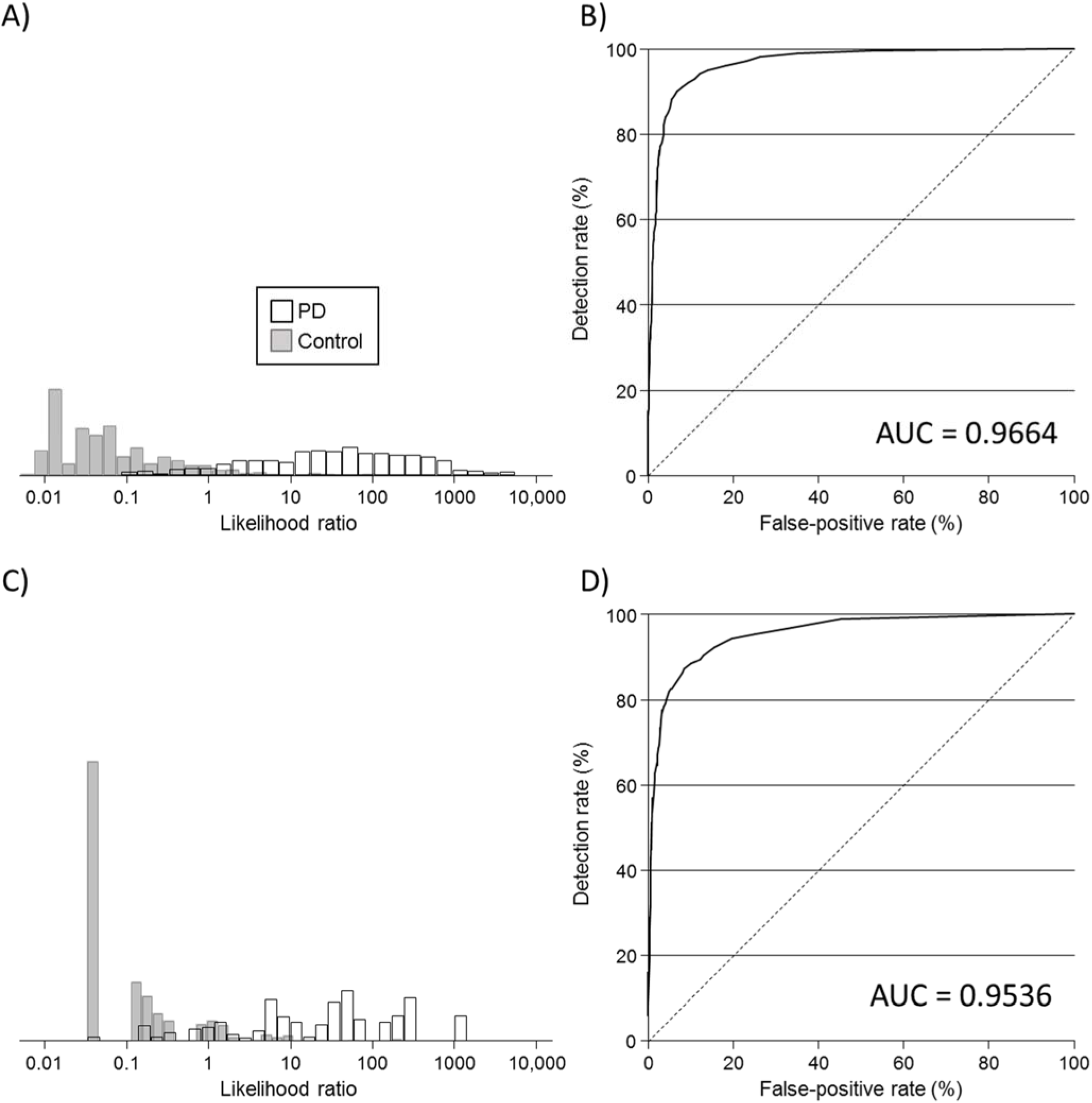
Distribution of likelihood ratios among Parkinson’s disease (PD) cases and controls, and the observed detection rate according to false-positive rate (receiver operating characteristic curve) for multivariate logistic regression models (see Table 1) based on all common odours to the UK and US versions of the UPSIT (A and B) and based on the 6 most odours most strongly associated with PD common to the UK and US versions of the UPSIT (C and D). AUC; area under the receiver operating characteristic curve

Supplementary Table 2 shows results for the multivariate model for the 6 odours previously identified as being predictive of hyposmia [Joseph et al 2019, Auger et al 2020]. Supplementary Figure 1A shows the distribution of likelihood ratios in PD patients and controls and Figure 1B shows the ROC curve based on these 6 odours. The median likelihood ratios were 6 in PD patients and 0.14 in controls (p<0.001) and ranged from 0.14 to 112 in PD patients and from 0.14 to 195 in controls. False-positive rates for 50, 60, 70 and 80% detection rates were 4.1%, 5.3%, 8.5% and 14.7% respectively; higher rates than for the 6 odours that were most strongly associated with PD. Corresponding internally validated estimates were 4.2%, 5.4%, 8.7% and 15.0%. The AUC was 0.88 and the internally validated AUC was also 0.88.

For the alternative approach examining the total UPSIT scores there was a small but significant decrease in UPSIT scores with increasing age (0.61 per 5 years of age, 95% CI 0.31 to 0.92; p<0.001) and males overall had lower scores than females (1.26 points lower, 95% CI 0.66-1.87; p<0.001). The regression equations for males and females were

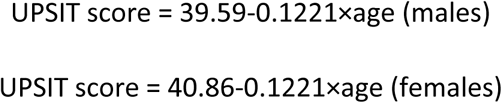

Using these equations to convert to delta values gave the distribution of scores shown in Supplementary Figure 2A. In both PD patients and controls, there was evidence of a mixture of two normal distributions. In PD patients the mixing proportions were 72% (*p*_*PD,1*_) following a distribution with mean -15.57 (*µ*_*PD,1*_) and standard deviation 4.17 (*σ*_*PD,1*_) and 28% (*p*_*PD,2*_) folllowing a distribution with mean -4.51 (*µ*_*PD,2*_) and standard deviation 4.35 (*σ*_*PD,2*_). In unaffected controls, the mixing proportions were 17% (*p*_*C,1*_) folllowing a distribution with mean 7.07 (*µ*_*C,1*_) and standard deviation 5.97 (*σ*_*C,1*_) and 83% (*p*_*C,2*_) folllowing a distribution with mean 0.50 (*µ*_*C,2*_) and standard deviation 2.81 (*σ*_*C,2*_). Using these values, the formula for calculating the likelihood ratio for specific UPSIT delta scores is:

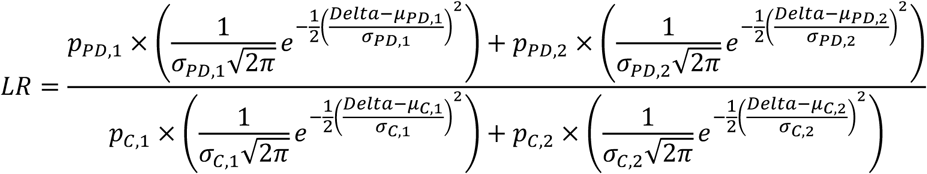

Supplementary Figure 2B shows the likelihood ratio according to UPSIT score MoM values. Risk reversal occurred below UPSIT delta values of -23.71 and above 4.15, and truncation limits were applied at these points. The median likelihood ratio in PD patients was 10.2 and in controls was 0.12 (p<0.001) with a range in likelihood ratios from 0.07 to 45. Supplementary Figure 2C shows an ROC curve for the UPSIT delta values. For observed detection rates of 50, 60, 70 and 80%, the false-positive rates were 2.5%, 3.6%, 4.7% and 8.7% respectively; higher than the false-positive rates derived from the results of the logistic regression analyses. The AUC was 0.74, lower than the AUC values based on the logistic regression analyses.

### Probable REM sleep behaviour disorder

Table 2 shows the results of the multivariate logistic regression analyses on items of the RBDSQ. Six of the 12 questions were significantly associated with being a PD case (Supplementary Table 3 shows univariate odds ratios for each question of the RBDSQ). Likelihood ratios are calculated by using the coefficients in Table 2 to generate the log odds, exponentiating and then dividing by (875/1314). Figure 2A shows the distribution of likelihood ratios and Figure 2B an ROC curve based on the results in Table 2. The median likelihood ratio was in 1.10 PD patients and 0.80 in controls (p<0.001) and the range in 0.34 to 69 PD patients and 0.34 to 29 in controls. False-positive rates for 30, 40, 50 and 60% detection rates were 4.0%, 7.2%, 12.2% and 20.2% respectively. Corresponding internally validated estimates were 4.0%, 7.2%, 12.3% and 20.3%. The AUC was 0.74, and the internally validated AUC was also 0.74.

**Table 2:** Results of multivariate logistic regression analyses of the questions that make up the REM sleep behaviour screening questionnaire (RBDSQ).

| RBDSQ question | Coefficient | OR (95% CI) | p-value | Likelihood ratio test statistics<br>between nested models |  |
| --- | --- | --- | --- | --- | --- |
| | | | | $\chi^2$ | p-value |
| 5. It thereby happened that I (almost) hurt my bed partner or myself | 1.566722 | 4.79 (3.26 to 7.04) | $1.44 \times 10^{-15}$ | 23.15 | $2.88 \times 10^{-52}$ |
| 3. The dream contents mostly match my nocturnal behaviour | 1.044969 | 2.84 (2.14 to 3.78) | $5.38 \times 10^{-13}$ | 73.62 | $9.47 \times 10^{-18}$ |
| 1. I sometimes have very vivid dreams | -0.872100 | 0.42 (0.34 to 0.51) | $5.89 \times 10^{-17}$ | 49.64 | $1.84 \times 10^{-12}$ |
| 6. I have or had any of the following phenomena during my dreams: |  |  |  |  |  |
| 6.1 Speaking, shouting, laughing very loudly | 0.7731019 | 2.17 (1.71 to 2.74) | $9.31 \times 10^{-11}$ | 57.56 | $3.28 \times 10^{-14}$ |
| 6.4 Things that fell down around the bed, e.g. bedside lap, book, glasses | 0.7478248 | 2.11 (1.38 to 3.24) | 0.0006 | 13.99 | 0.0002 |
| 4. I know that my arms or legs move when I sleep | 0.3166089 | 1.37 (1.09 to 1.73) | 0.0076 | 7.03 | 0.0080 |
| Constant | -0.6268203 | 0.53 (0.46 to 0.62) | $1.50 \times 10^{-17}$ | | |

**Figure 2:**
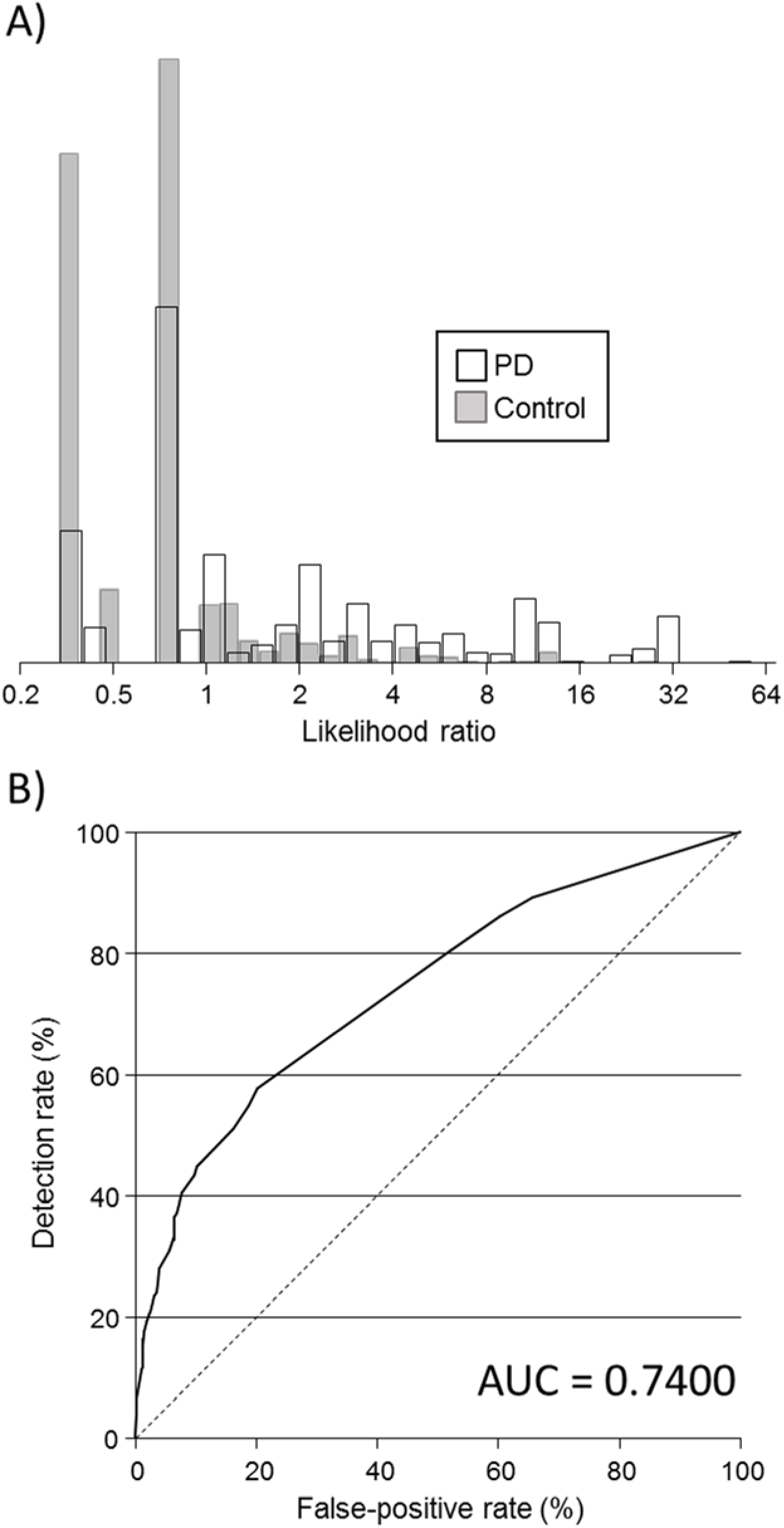
Distribution of likelihood ratios among Parkinson’s disease (PD) cases and controls (A), and the observed detection rate according to false-positive rate (receiver operating characteristic curve; B) for the multivariate logistic regression model based on questions from the REM sleep behaviour screening questionnaire (see Table 2). AUC; area under the receiver operating characteristic curve

For the alternative approach examining the RBDSQ scores, Supplementary Figure 3A shows the percentage of participants with each RBDSQ score. Neither age nor gender influenced scores so adjustment was not necessary. Figure 3B shows an ROC curve for the RBDSQ. Screening performance was modest, for example, when we used an RBDSQ score of 5 or more the detection rate was 35% and the false-positive rate was 15%. Using a lower RBDSQ cut-off score of 4 the detection rate increases to 45% and the false-positive rate to 23%. The AUC was 0.6305. Figure 3C shows the likelihood ratio according to score together with a regression line. The likelihood ratios range from 0.70 for a score of 0 to 23.19 for a score of 12 and are given by the following formula;

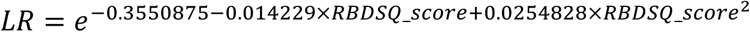

**Figure 3:**
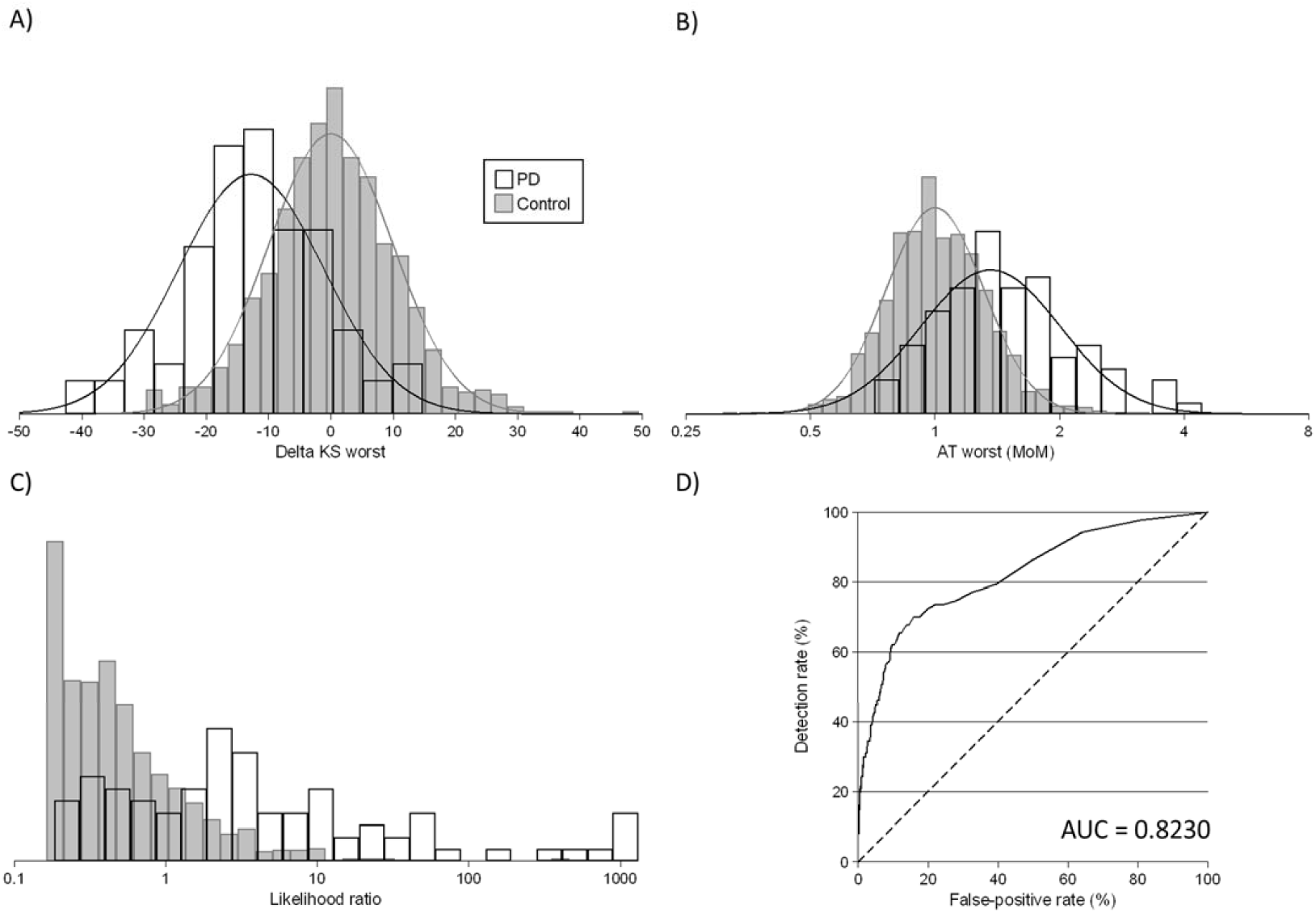
Distributions of BRAIN test parameters kinesia score (delta KS; A) and akinesia time (multiple of the median [MoM] AT; B) in Parkinson’s disease (PD) patients and controls, distribution of likelihood ratios based on the multivariate Gaussian distributions of KS and AT (C) and the observed detection rate according to false-positive rate (receiver operating characteristic curve) based on the multivariate Gaussian distributions of KS and AT (D) AUC; area under the receiver operating characteristic curve

Calculating likelihood ratios according to RBDSQ scores performed less well in predicting PD than basing likelihood ratios on the logistic regression approach; the AUC was 0.63.

### Quantitative motor impairment

For the BRAIN test scores, KS followed a Gaussian distribution while AT was log-transformed. There was a small but significant decrease in KS scores with increasing age (-1.02 per 5 years of age, 95% CI -0.31 to -1.72; p=0.005) and females overall had higher scores (1.49 higher, 95% CI 0.10-2.87; p=0.035). The regression equations were

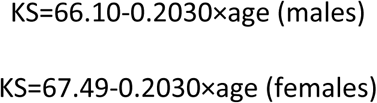

There was a small but significant increase in log (natural) AT scores with increasing age (0.04 per 5 years of age, 95% CI 0.01 to 0.06; p=0.001) and females overall had higher scores (0.08 higher, 95% CI 0.04 to 0.12; p<0.001). The regression equations were

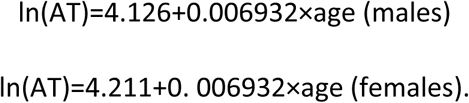

After transforming values into delta values for KS, and MoM values for AT using the above regression equations, mean KS values were 12.8 points lower in PD patients than in controls, and AT values 36% higher (both p<0.001). Supplementary Figure 4 shows probability plots for delta KS values (A) and AT MoM values (B), with AT MoM values plotted on a log scale. Delta KS values start to deviate or the data were sparse below -30 and above 10 and for AT below 0.5 MoM and above 3.0 MoM. These values were therefore used as truncation limits. However, the point of risk reversal for AT was at 0.747 so values less than this were truncated. All distributions were reasonably Gaussian as indicated by the points roughly falling on straight lines. Figure 3 shows the distribution of delta KS (A) and AT MoM (B) values in PD patients and controls. KS had the best discrimination between PD patients and controls. Supplementary Table 4 shows the parameters (means, standard deviations, correlation coefficients, truncation limits) in PD patients and controls. Likelihood ratios are given by the following formula

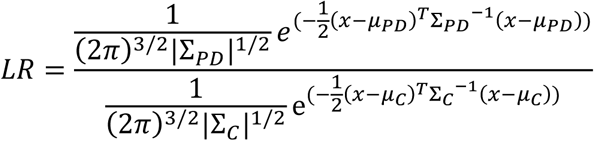

Where *Σ*_*PD*_ is the covariance matrix in PD patients, |*Σ*_*PD*_| is the determinant of the covariance matrix in PD patients, *µ*_*PD*_ is the vector of the means of KS, AT and IS in PD patients, *Σ*_*C*_ is the covariance matrix in controls, |*Σ*_*C*_| is the determinant of the covariance matrix in controls, *µ*_*C*_ is the 1×2 vector of the means of KS MoM values and ln(AT MoM values) in controls, and *x* is the 1×2 vector of MoM values (KS MoM, ln(AT MoM)); *T* stands for transpose. Figure 3C shows the distributions of likelihood ratios in PD patients and controls. The median likelihood ratio in PD patients was 2.73 and in controls was 0.39 (p<0.001) with a range in likelihood ratios from 0.18 to 1311 in PD patients and 0.16 to 458 in controls. Figure 3D shows an ROC curve for the combination of delta KS and AT MoM values. False-positive rates for 30, 40, 50 and 60% detection rates were 1.8%, 4.0%, 6.6% and 20.2% respectively. Corresponding internally validated estimates were the same to one decimal place. The AUC was 0.82 and the internally validated AUC was the same to two decimal places.

## DISCUSSION

Our results show that maximising information on markers for PD that are either continuous or discrete substantially extends the range of likelihood ratios than is achieved by dichotomisation. In comparison to dichotomous likelihood ratios for olfactory performance of 6.4 for those with hyposmia versus 0.40 for those with normosmia^7^, our olfactory likelihood ratios ranged from 45 to 0.07 based on the full range of olfactory performance (using the total score from the 40-item UPSIT), and ranged from 0.009 to about 5500 using the logistic regression approach that identified 16 odours that were significantly associated with PD. Similarly, RBD likelihood ratios ranged from 0.70 for a total RBDSQ score of 0, through to 23.19 for a score of 12, or from 0.34 to 29 using the logistic regression approach. Both approaches comparing favourably to likelihood ratios of 2.8 for those with probable RBD and 0.89 for those without probable RBD previously reported^7^, with the logistic regression approach being superior Finally, likelihood ratios for finger tapping speed based on two BRAIN test parameters of between about 0.16 and 1300 can be achieved instead of dichotomising compared to likelihood ratios of 3.5 for those with abnormal quantitative motor testing and 0.60 to those without.^7^

The logistic regression approach to UPSIT and RBD provides more information for examining total UPSIT and RBDSQ scores, and offer an improvement to simply dichotomising to scores. There are a number of different tests of smell on the market, such as the Sniffin’ Sticks and conversion of scores from this test to the full 40-item UPSIT has been described.^21^ For researchers using alternative smell tests, conversion could be performed and likelihood ratios for the full UPSIT applied (Supplementary Figure 2B). Otherwise a logistic regression approach could be used given sufficient data. We presented two 6-item odour tests, one from previous work that selected items on their ability to predict hyposmia in healthy controls^13,19^, and a new selection based on their ability to different PD cases from controls. The 6-item test reported here had similar performance to the test using 16 of the 32 odours common to both the UK and US UPSIT. The 6-item test based on previous work performed less well, but this test may have an advantage in that it was primarily designed to test for hyposmia, in which case it could be more generalizable to diseases other than PD. In any case a 6-item test could offer substantial financial savings over routinely using the full UPSIT.

Dichotomising continuous variables comes at cost of loss of information^15^, and is avoided in other areas of medical risk assessment calculations (e.g. using actual blood pressure readings rather than dichotomising as hypertensive or not above a fixed value). In this study, we have used an approach similar to that which has been used for many years in prenatal screening for Down syndrome, as well as more recently in prenatal screening for trisomy 18, trisomy 13 and preeclampsia. In the same way as MoM values take account of natural changes in ultrasound and serum markers with gestational age in prenatal screening for Down syndrome and similar conditions^22,23^, our delta and MoM values take account of normal changes in smell loss and tapping parameters with age, and also between males and females. Researchers wishing to use delta or MoM values could generate these from their own data in the same way as done here, using regression analysis among those without PD, or in a cohort study by regression analysis in all participants. Emerging blood or other biomarkers for PD would also benefit from the use of delta or MoM values instead of mass units, with the added advantage of accounting for of systematic differences between different assays and laboratories by calculating delta or MoM values based on local data. The use of multivariate Gaussian distributions also allows for a modular approach i.e. adding new markers as they are discovered, without needing data on each marker to be measured in the same participants.

A weakness of this study is that the estimates of screening performance are based on the marker distribution in those with diagnosed PD. In practice such risk estimation will take place before diagnosis, so screening performance estimates in a ‘healthy population’ are likely to be lower than those presented here. We have however examined the likelihood ratios presented here for smell, tapping speed and RBDSQ scores together with age and other factors, such as smoking status and family history of PD, to determine by how much the spread of risk increases in the PREDICT-PD cohort, and will report this separately. It is recognised that estimates of performance of a predictive model are overestimated when determined on the sample from which the model is derived^20^ but our internally validated estimates of performance were very similar. However, the approach used here would also need to be validated in independent datasets. A further weakness of this study is that data on PD patients came from a different source to that of controls. Although the age and gender profiles were similar between PD patients and controls we cannot exclude the presence of residual confounding, reinforcing the need for external validation.

In summary this study shows that maximising information on continuous and discrete markers for PD has potential to improve the ability of algorithms to detect PD and this study provides the methods for incorporating this approach into other algorithms.

## Data Availability

It is the intention of the authors to make study data available for sharing

## ACKNOWLEDGMENTS

We would like to thank all the participants in the PREDICT-PD pilot cohort and the participants in the Tracking Parkinson’s study

## AUTHORS’ ROLES

1) Research project: A. Conception, B. Organization, C. Execution; 2) Statistical Analysis: A. Design, B. Execution, C. Review and Critique; 3) Manuscript: A. Writing of the first draft, B. Review and Critique.

J.P.B.: 1A, 1B, 1C, 2A, 2B, 2C, 3A, 3B

S.D.A.: 2A, 2B, 2C, 3B

A.E.S.: 1A, 1B, 1C, 2C, 3B

D.G.G.: 1A, 1B, 3B

S.K.: 1A, 1B, 3B

G.G.: 1A, 1B, 3B

A.J.L.: 1A, 1B, 3B

J.C.: 2C, 3B

A.J.N.: 1A, 1B, 1C, 2C, 3B

